# Life-course patterns of heavy menstrual bleeding and menstrual pain across multiple population-based cohorts

**DOI:** 10.64898/2026.10.07.26364868

**Authors:** Sampurna Kundu, Gemma Sawyer, Maria Christine Magnus, Laura D Howe, Abigail Fraser, Martha Hickey, Heather O’Mahen, Jessica Tyrrell, Jacqueline A. Maybin, Hilary Critchley, Dani J. Barrington, Gemma C Sharp

## Abstract

**Background:** Heavy menstrual bleeding (HMB) and menstrual pain (MP) are among the most common menstrual cycle related symptoms with substantial impacts on quality of life. Despite this, population-level evidence on their prevalence across the life course is scarce.

**Methods:** We conducted descriptive analyses of six longitudinal population-based cohorts from the United Kingdom, United States, Norway, and Australia. We estimated cumulative prevalence (across all available timepoints) and age-specific point prevalence of HMB and MP. Cumulative prevalence estimates were stratified by sociodemographic factors, where data permitted. We also examined HMB and MP incidence in adolescent cohorts with repeated measurements.

**Findings:** HMB and MP are commonly reported, with cumulative prevalence across all the cohorts and timepoints (i.e., ever reported) ranging from 36% to 89% for HMB and 46% to 72% for MP. In adolescence, MP (point-prevalence: 33% to 47%) is more commonly reported than HMB (16% to 24%), whereas in later ages (45 years and above), HMB (47% to 70%) is more prevalent than MP (24% to 40%). We found no consistent evidence that symptoms were more likely to co-occur than occur independently; patterns varied across life stages and cohorts. Symptoms were more common in those from lower socioeconomic backgrounds and minoritised ethnic groups. Incidence of both HMP and MP peaked in the years immediately following menarche.

**Interpretation:** Most women experience heavy and/or painful periods at some point. MP is more common in adolescence and HMB in later reproductive life. Generating robust life-course evidence on menstrual symptom patterns is essential to inform clinical guidelines, reduce diagnostic delays, and address inequalities in menstrual health.

## Introduction

There is growing recognition of the impact of heavy and painful menstrual periods on health ^1^, quality of life ^2^, education ^3^, and employment ^4^. Heavy menstrual bleeding (HMB) is defined by NICE (National Institute for Health and Care Excellence) as a volume of blood loss that interferes with an individual’s physical, social, emotional, and/or material quality of life ^5^. This was updated in 2007 from an earlier objective definition (>80ml) that was difficult to assess and did not correspond with clinical outcomes ^6^. Dysmenorrhea (referred to as menstrual pain (MP) in this paper) is also defined subjectively as painful cramping, usually in the lower abdomen, occurring shortly before and/or during menstruation ^7^. HMB and MP may occur independently or together, either in the absence of any identifiable pathology, or due to conditions such as endometriosis, adenomyosis, or uterine fibroids (leiomyoma) ^8,9^.

HMB and MP prevalence estimates are highly variable. A recent systematic review of population-based studies conducted across 20 high- and low-middle-income countries (HICs and LMICs) reported HMB prevalence between 15.2-65% ^10^. Similarly, a review of community-based studies from 11 HICs and LMICs found MP prevalence estimates ranging from 6-91% among women of reproductive age ^11^. This variation could reflect true differences across the life course, sociodemographic groups, and geographical settings, but likely also reflects methodological differences in how these symptoms are defined and measured, as well as the tendency for menstrual symptoms to be under-reported, normalised, and dismissed in general ^12^. Existing estimates are largely drawn from cross-sectional studies, thus limiting the understanding of how patterns change overtime. Furthermore, HMB and MP frequently co-occur ^11^; however, there are no population-based studies reporting estimates of this co-occurrence.

More reliable prevalence estimates are needed to understand the population burden of menstrual health symptoms and how they vary across age, population, and sociodemographic groups. Health records and registries are unlikely to provide accurate prevalence estimates due to low healthcare-seeking for these symptoms and poor documentation when care is sought. In a 2012 cross-sectional general population survey of >4,500 women across five European countries, 46% of those with HMB (n=1,225) had never consulted a clinician about their symptoms ^13^. Population-based longitudinal cohorts provide valuable data on menstrual symptoms, including investigation of how prevalence changes over time to identify life stages when symptoms are most common.

This present study brings together six cohort studies from multiple countries that have collected data on HMB and MP. We leverage prospective and longitudinal data to describe the prevalence of self-reported HMB and MP across the life course, including by sociodemographic groups, and to examine symptom onset following menarche.

## Methods

We followed the STROBE (Strengthening the Reporting of Observational Studies in Epidemiology) guidelines in the reporting of this study (Supplementary File 1).

### Cohorts

We used data from six cohorts: the Adolescent Brain Cognitive Development Study (ABCD; United States, 9-10-year olds in 2016-2018) ^14^, the Avon Longitudinal Study of Parents and Children (ALSPAC; United Kingdom, pregnant women/Generation 0 [G0] in 1991-1992 and their offspring/Generation 1 [G1]) ^15,16^, the Raine Study (Western Australia, births/Generation 2 [Gen2] in 1989-1991) ^17^, the Norwegian Mother, Father and Child Cohort Study (MoBa; pregnancies in 1999-2008) ^18^, and the 1970 British Cohort Study (BCS70; United Kingdom, births in a single week in April 1970) ^19^. These cohorts were selected because they provide repeated measures of HMB and MP and, together, span early adolescence to midlife (Figure 1). More information is provided in Supplementary File 1.

**Figure 1:**
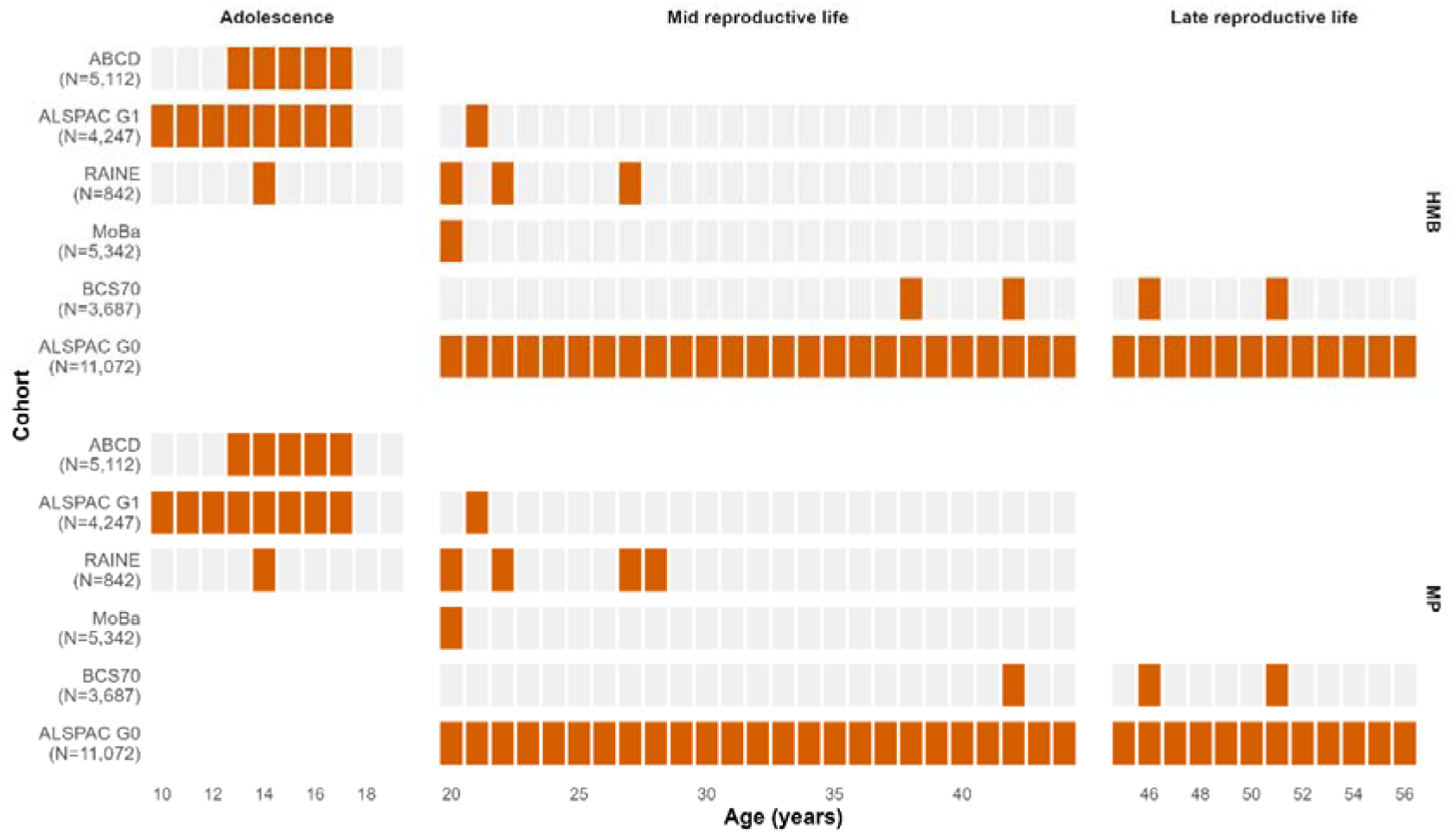
Age coverage (timepoints) of heavy menstrual bleeding (HMB) and menstrual problems (MP) by life stages across cohorts.

### Participants

We included female participants with available data on HMB or MP at one or more assessment timepoints within each cohort (see Supplementary Figure S1 for flowchart). Figure 1 summarises sample sizes and the timing of menstrual symptom assessments (ABCD: N=5,112; ALSPAC G1: N=4,247; the Raine Study: N=842; MoBa: N=5,342; BCS70: N=3,687; ALSPAC G0: N=11,072).

### Data harmonization

#### Menstrual symptoms

Menstrual symptom questions varied across cohorts and timepoints in terms of wording, indicators, and symptom reporting time range (Table 1). To facilitate cross-cohort comparisons, we derived binary variables of any HMB (HMB with or without MP versus no HMB) and any MP (MP with or without HMB versus no MP) using cohort-specific measures (Supplementary Tables S1a-f).

**Table 1.** Summary of HMB and MP indicators assessed in each cohort.

| Cohort | Heavy menstrual bleeding |  | Menstrual pain |  | Reporting range |
| --- | --- | --- | --- | --- | --- |
|  | Indicator | Timepoint(s) | Indicator | Timepoint(s) |  |
| ABCD | Self-reported 'heavy menstrual flow' | 13-17 | Severity ≥6 rated on 0-10 VAS* | 13-17 | On average |
| ALSPAC G0 | Self-reported 'very'/'moderately' heavy flow | 20-56** | Self-reported 'very'/'moderately' painful periods | 20-56** | Most recent periods |
| ALSPAC G1 | Self-reported 'heavy or prolonged bleeding'. | 10-17 | Self-reported 'severe period cramps' | 10-14, 16-17 | Ever experienced |
|  |  |  | Self-reported 'severe'/'moderate' pain with period | 15 |  |
|  | Self-reported 'very'/'moderately heavy periods | 21 | Self-reported 'very'/'moderately painful periods | 21 | Most recent periods (if stopped, periods before they stopped) |
| Raine | Self-reported 'heavy menstrual periods' | 14 | Self-reported 'painful menstrual cramps' | 14 | 14 – Not specified<br>20-22 – Regular/Common experience<br>27 – Past month<br>28 – Last 12 months |
|  | Double protection OR flooding OR super protection and >4 changes on heaviest day | 20-22 | Severity ≥6 rated on 0-10 VAS | 20-28 |  |
|  | Self-reported 'heavy'/'very heavy' flow OR >4 soaked products on heaviest day | 27 |  |  |  |
| BCS70 | Doctor consultation for heavy periods in the last year | 38 | Self-reported 'painful periods' in the past four years | 42-46 | 38 – In last year<br>42-51 – Time since last interview or four years prior |
|  | Self-reported 'heavy periods' in the past four years | 42-46 | Self-reported 'painful periods' | 51 |  |
|  | Self-reported 'heavy periods' | 51 |  |  |  |
| MoBa | Changing of products >6 times a day OR changing every two hours OR bleeding through products OR double protection OR wake at night to change products OR blood on bedding | 20 | Self-reported 'lower abdominal pain during menstruation' | 20 | Usually |

\*VAS: Visual Analogue Scale (commonly used in pain assessment); \*\*Actual ages were used for ALSPAC G0

We also derived a four-category variable: (1) neither symptom, (2) HMB only, (3) MP only, and (4) both HMB and MP. Participants who did not answer menstrual symptom questions, had not started their menstruation, or had not experienced menstruation in the past 12 months at a particular follow-up timepoint were coded as missing for that timepoint.

#### Covariates

##### Age

In cohorts where participants were recruited at roughly similar age (all except for ALSPAC G0), we defined assessment age as mean participant age (in years) at each timepoint (or questionnaire). In ALSPAC G0, participants were recruited during pregnancy, meaning participant age varies more substantially than the other cohorts. Therefore, instead of analysing ALSPAC G0 data by timepoint (or questionnaire), we examined this by participant age at assessment. We calculated age at assessment from participant age at questionnaire completion (rounded down to the nearest year) and grouped responses at the same age year (meaning different timepoints/questionnaires contribute to the same age at assessment for different participants) (Supplementary File 1). Individual participant age at assessment was also utilised in ABCD and ALSPAC G1 to examine symptom onset (incidence) with chronological age (rounded down to the nearest year). Timepoint age was categorised into adolescence (<20 years, according to WHO definition of adolescence ^20^), mid-reproductive life (20 to 44 years), and late reproductive life (45+ years).

##### Gynaecological age

As menstrual cycles and symptoms may take time to stabilise following menarche, we examined whether incidence varied by time since menarche (or gynaecological age) ^21^. In cohorts with prospective data from menarche (ABCD and ALSPAC G1), we calculated gynaecological age by subtracting age at menarche (based on the first reported age at menarche across repeated assessments) from age at assessment (Supplementary File 1).

##### Childhood socioeconomic position (SEP)

Three measures of SEP were examined, categorised into Low, Middle, or High (although not every cohort had all three measures available). Consistent definitions between cohorts were used where possible; however, differences in questions, response options, and cohort timing meant this was not always possible and therefore categories may represent relative SEP within each cohort (Supplementary Table S2 for details). Variables were parent- (ALSPAC G1, BCS70, ABCD, MoBa, the Raine study) or self-reported (ALSPAC G0) at baseline or during childhood.

###### Parental Education

We derived highest maternal and paternal education in all cohorts, except ABCD where a single parental measure was available and harmonised into Low, Middle, and High (Supplementary Table S2).

###### Childhood Household Income

We harmonised household income during childhood into Low, Middle, and High based on cohort-specific income distributions (except ALSPAC where this was not available) (Supplementary Table S2).

###### Parental Occupation

As with education, we derived maternal and paternal occupational class during childhood in BCS70 and both ALSPAC generations into Low (partly skilled and unskilled), Middle (skilled), and High (professional and managerial/technical) (Supplementary Table S2).

##### Ethnicity

In most cohorts, we derived a binary ethnicity variable (White and Non-white) due to limited ethnic diversity. In ABCD, however, we derived a four-category ethnicity variable (White, Black, Asian, or Other) to reflect greater ethnic diversity. Ethnicity was not available in MoBa.

### Analysis

All statistical analyses were conducted using R (version 4.5.0), with data preparation performed in Stata (version 19.5). Analyses were performed separately within each cohort due to differences in study design, age coverage, and measurement.

#### Cumulative prevalence

We defined cumulative prevalence as the proportion of participants reporting any HMB, any MP, and either HMB/MP at least once across all timepoints (symptom co-occurrence was not examined). The total sample (denominator) was participants who contributed menstrual data at least once. We excluded MoBa from this analysis as only one timepoint with menstrual data is available, meaning cumulative prevalence is equivalent to point prevalence.

To examine sociodemographic variation in menstrual symptoms, crude prevalence ratios (PRs) comparing symptom prevalence across measures of SEP and ethnicity (where available) were calculated using the highest SEP category or White ethnicity as reference groups. Corresponding 95% confidence intervals (CIs) were calculated using the Wald method on the log-transformed PR.

#### Point prevalence

We calculated age-specific (point) prevalence at each available timepoint for any HMB, any MP, isolated, and co-occurring symptoms, with the denominator defined as participants contributing menstrual data at the corresponding timepoint. We excluded adolescent timepoints from ALSPAC G1 because participants were asked to report whether they had *‘ever’* experienced a symptom, meaning the reporting range was not specific to the assessment timepoint (Table 1). We report the 95% CIs for all prevalence estimates, calculated using the Wilson score method for binomial proportions.

#### Incidence in adolescence

In cohorts with prospective data from menarche (ABCD and ALSPAC G1), we estimated symptom incidence (any HMB and any MP) according to both chronological and gynaecological age. Incident cases were defined as the first report of a symptom (i.e., not reported that symptom at earlier assessments following menarche). At each age, the denominator comprised participants contributing menstrual data who remained at risk (i.e., had not previously reported the symptom). 95% CIs were calculated using the Wilson score method for binomial proportions.

## Results

### Sample characteristics

Sociodemographic characteristics of the analytical samples are presented in Table 2. Cohorts were predominantly White, although ABCD has greater ethnic diversity than other cohorts. SEP variables also varied across cohorts, with contemporary cohorts (ABCD and MoBa) generally having higher levels of parental education and household income than older cohorts (i.e., BCS70 and ALSPAC G0).

**Table 2.**
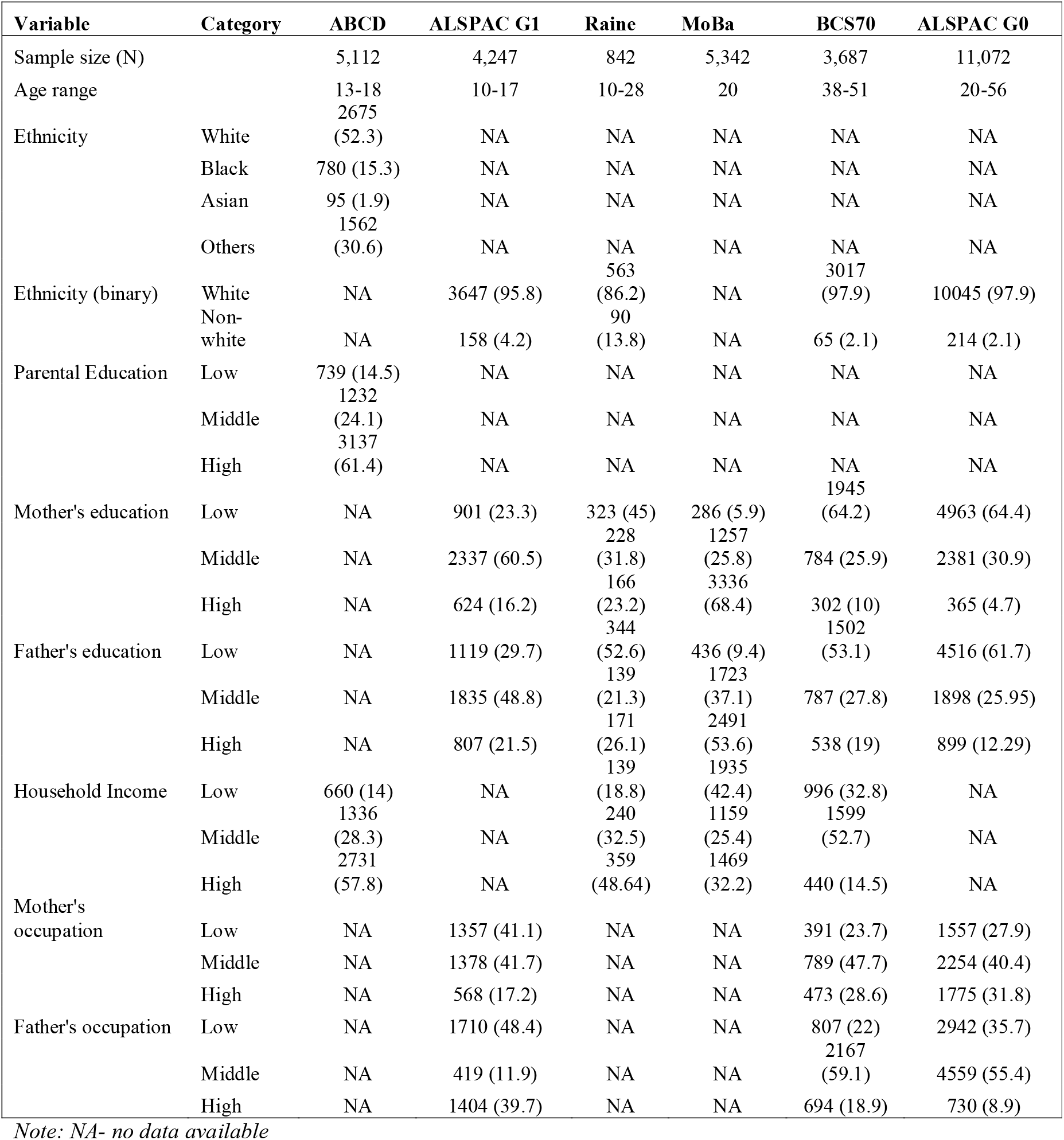
Sociodemographic characteristics of the analytical samples across cohorts (n (%)).

### Cumulative prevalence

Across all cohorts, a high proportion of participants reported experiencing menstrual symptoms at least once across available assessments (Figure 2; full results in Supplementary Table S3). For HMB, cumulative prevalence ranged from 36% (95% CI 35-38%) (ABCD) to 89% (95% CI 88-90%) (ALSPAC G0), while for MP, it ranged from 46% (95% CI 44-47%) (BCS70) to 72% (95% CI 70-73%) (ALSPAC G1). MP was more common than HMB in adolescence/young adulthood, whereas HMB was more common in later reproductive life.

**Figure 2:**
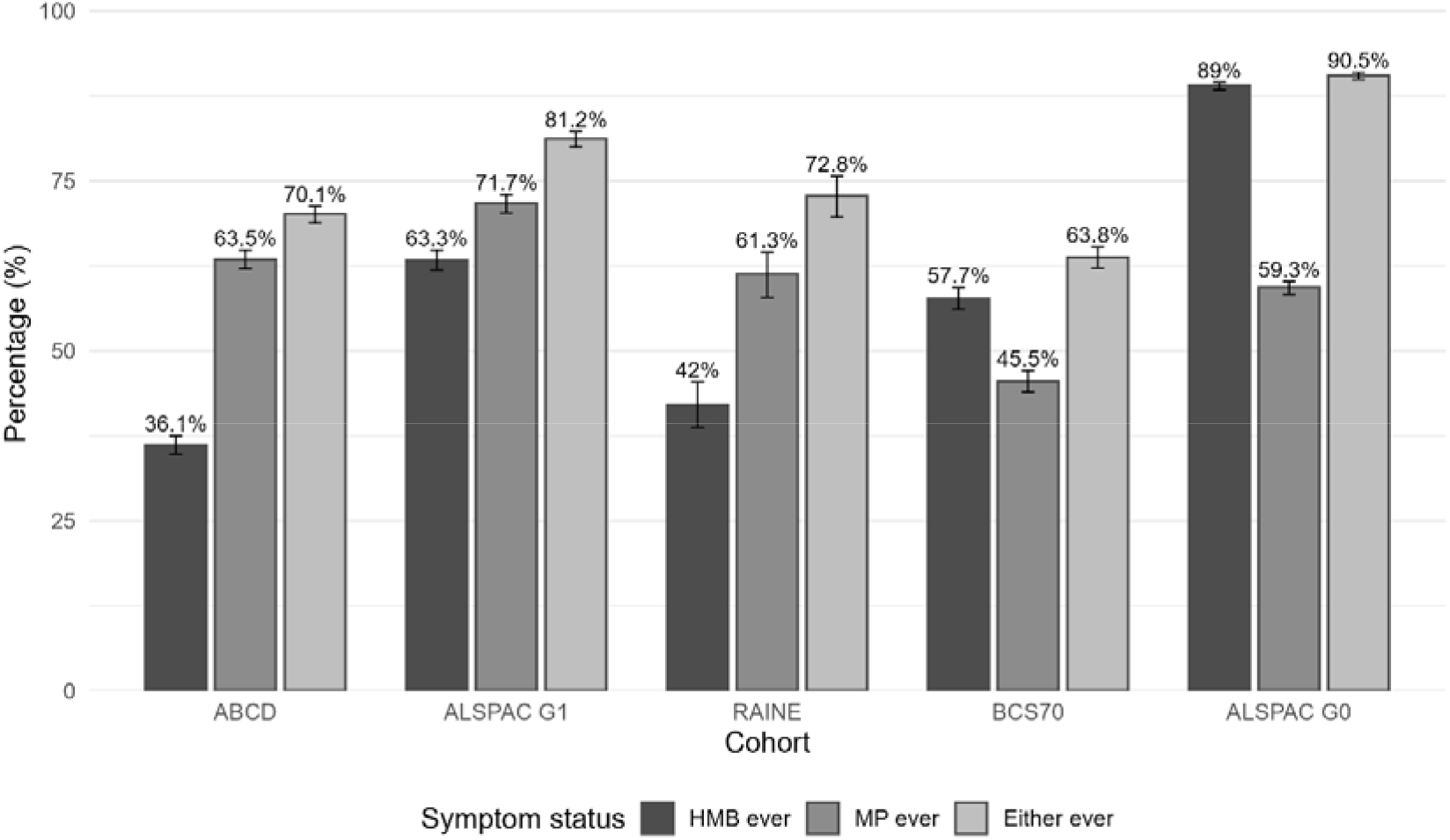
Cumulative prevalence of HMB, MP, and either HMB or MP across cohorts.

Differences between cohorts likely reflect variation in age coverage and duration of follow-up, with higher cumulative prevalence generally observed in cohorts with longer follow-up into adulthood.

#### Sociodemographic variation in cumulative prevalence

We observed evidence that cumulative prevalence of menstrual symptoms was generally higher among women with lower SEP factors, although differences varied by cohort (Supplementary Figures S3-S4 & Table S4). This was most evident in ABCD where the cumulative prevalence of either HMB or MP was 78% versus 65% in the lowest compared to highest income group (prevalence ratio PR 1.20; 95% CI 1.14-1.26) and the cumulative prevalence of MP was 73.9% versus 57.0% (PR 1.30; 95% CI 1.22, 1.37), respectively.

Similarly, in BCS70, MP (PR 1.19; 95% CI 1.04-1.36) and either HMB or MP (PR 1.12; 95% CI 1.02-1.23) were more prevalent in the lowest compared to highest maternal occupation group. Although differences according to SEP were weaker in other cohorts, with small magnitude PRs and/or CIs overlapping the null, results generally supported the same patterns.

Ethnic differences were most apparent in ABCD, reflecting its greater diversity (Supplementary Figure S5 & Table S4). Black participants reported the highest prevalence of MP (75.1%; PR 1.27; 95% CI 1.20-1.33 compared with White participants) and either symptom (79.5%; PR 1.20; 95% CI 1.15-1.25), whereas Asian participants had the lowest prevalence across outcomes. In remaining cohorts (except the Raine Study), prevalence tended to be somewhat higher for Non-white compared to White participants, although differences were small.

### Point prevalence

In ABCD and the Raine Study (adolescent cohorts; aged < 20), MP was consistently more common than HMB, with the point prevalence of any MP ranging from 33-47% in ABCD (compared to 16-24% for any HMB) and 37% in Raine (26% for any HMB) (Supplementary Table S5). In ABCD, which repeatedly assessed menstrual symptoms across adolescence, the point prevalence of MP and HMB tended to increase with chronological age; however, this was more marked for MP (Figure 3). During adolescence, MP most often occurred in isolation, whereas HMB tended to occur alongside MP instead of in isolation.

**Figure 3:**
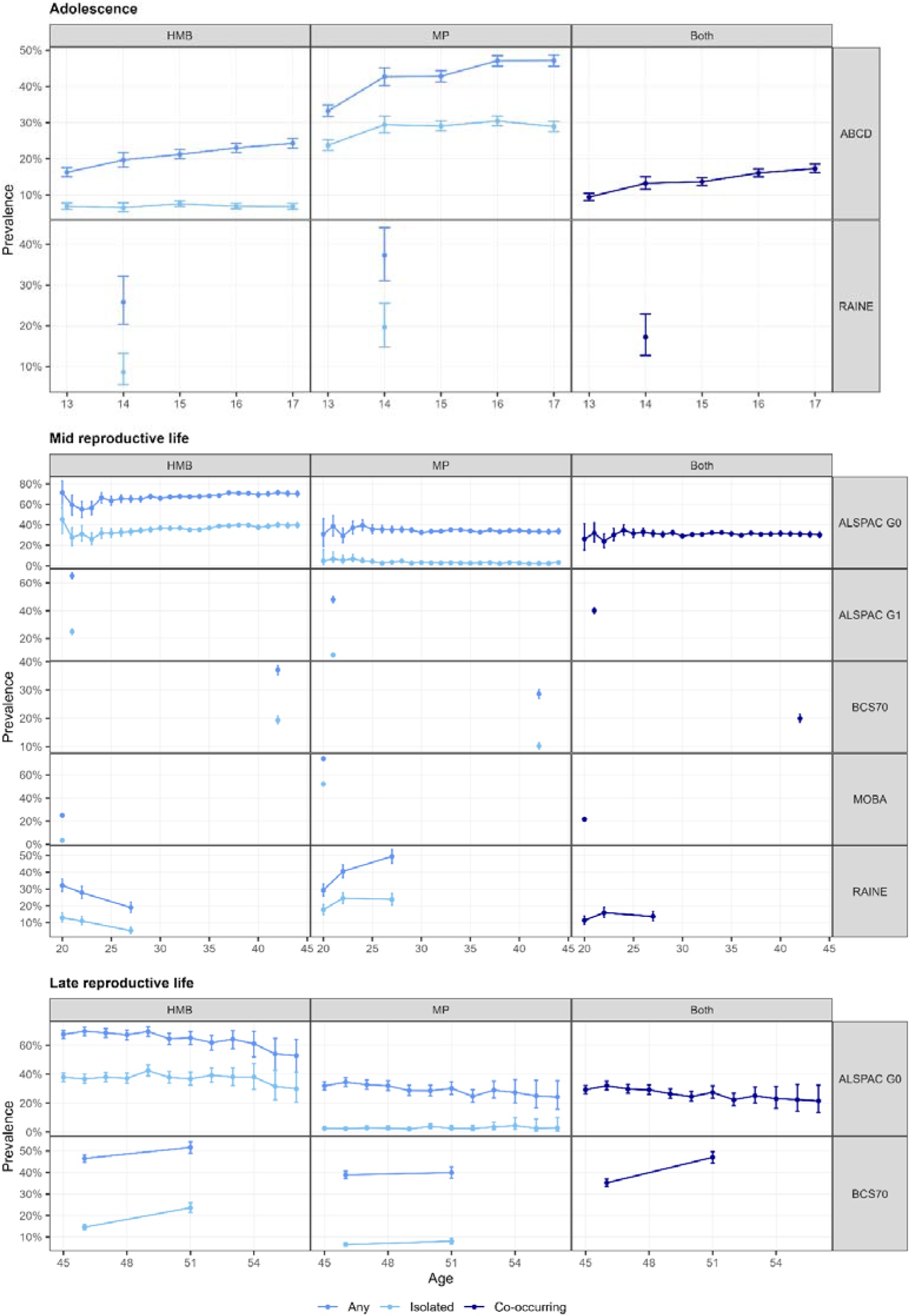
Age-specific point prevalence of HMB, MP and co-occurrence of both across cohorts.

In cohorts with mid reproductive aged participants (aged 20-44), no consistent trends were observed across more than two cohorts (Figure 3). In MoBa and Raine, MP (MoBa 74%; Raine 29-49%) was more prevalent, and primarily occurred in isolation, than HMB (MoBa 25%; Raine 19-32%). In contrast, in ALSPAC G0 and BCS70, HMB (ALSPAC G0 55-71%; BCS70 37-77%) was consistently more prevalent than MP (ALSPAC G0 29-40%; BCS70 29%) (Supplementary Table S5). HMB occurred alongside MP and in isolation at similar rates, whereas MP primarily occurred alongside HMB. In cohorts with more than one timepoint (ALSPAC G0 and Raine), we did not see consistent trends by age: in ALSPAC G0, prevalence of both symptoms was mostly stable, whereas in Raine the prevalence of HMB decreased from age 20 to 27 while MP increased over the same period.

In cohorts with late reproductive aged participants (aged ≥45; ALSPAC G0 and BCS70), HMB was more prevalent than MP, and only 2-10% of participants in both cohorts reported isolated MP (Supplementary Table S5). In BCS70, HMB prevalence increased between age 46 and 51 (47% to 52%), while MP prevalence was relatively stable (39-40%). In contrast, prevalence estimates in ALSPAC G0 decreased somewhat with increasing age (Figure 3).

### Incidence in adolescence

In ABCD and ALSPAC G1 (cohorts with repeated measures of menstrual symptoms from menarche), incident MP was consistently higher than incident HMB. However, we found marked differences in patterns by chronological age between cohorts (Figure 4). In ABCD, HMB incidence was relatively stable across most of adolescence (starting from 17% at age 11 gradually dropping to 9% at age 17) whereas MP incidence peaked at age 13 (36%). In ALSPAC G1, however, incidence of both symptoms tended to be lower at earlier ages (e.g., 18% for MP and 26% for HMB at age 11-12), increasing to a peak around age 14-15 (49% for MP at 15; 29% for HMB at 14), before declining thereafter (Supplementary Table S6).

**Figure 4:**
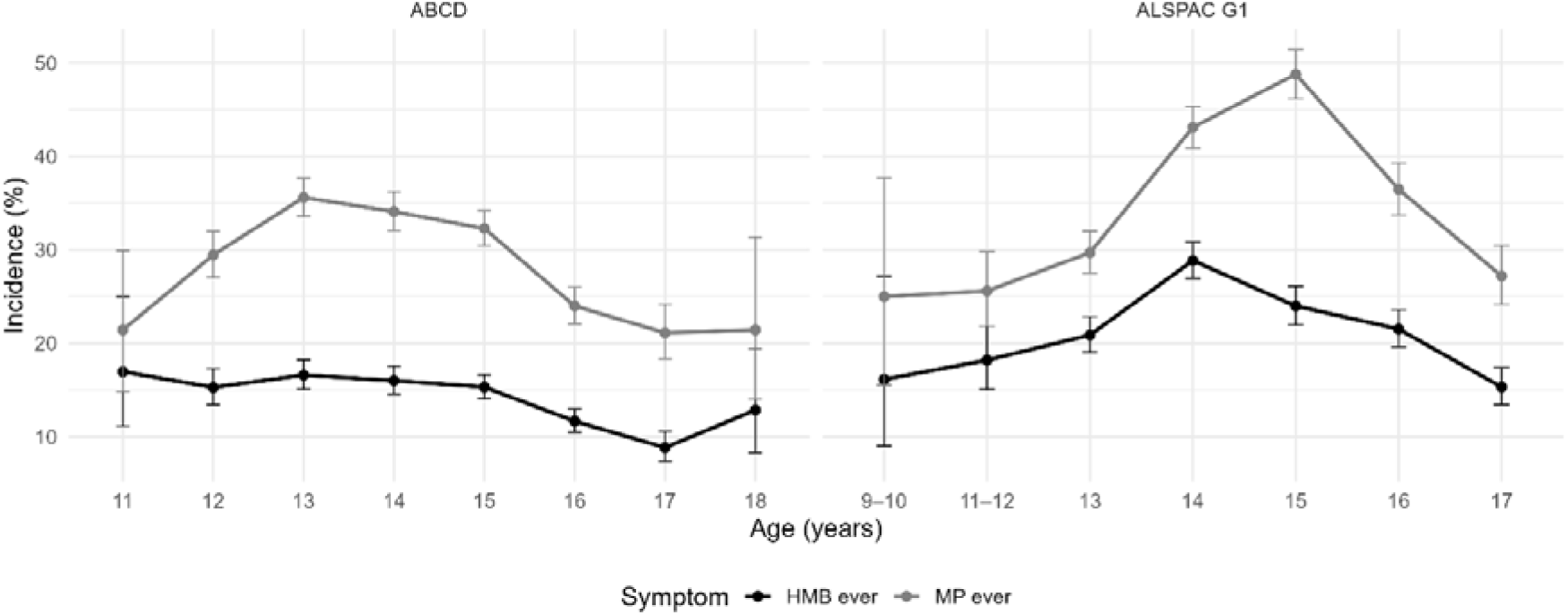
Incidence of heavy menstrual bleeding (HMB) and menstrual pain (MP) by age across adolescent and early adulthood cohorts.

When we examined incidence by gynaecological age (Figure 5), MP incidence remained consistently higher than HMB incidence. Incidence of both symptoms rose and peaked in the first few years following menarche. In ABCD, incidence of both symptoms peaked 2 to 3 years following menarche, whereas, in ALSPAC G1, HMB peaked earlier following menarche (0.5 to 1 year) and MP peaked slightly later (3 years post-menarche) (Supplementary Table S6).

**Figure 5:**
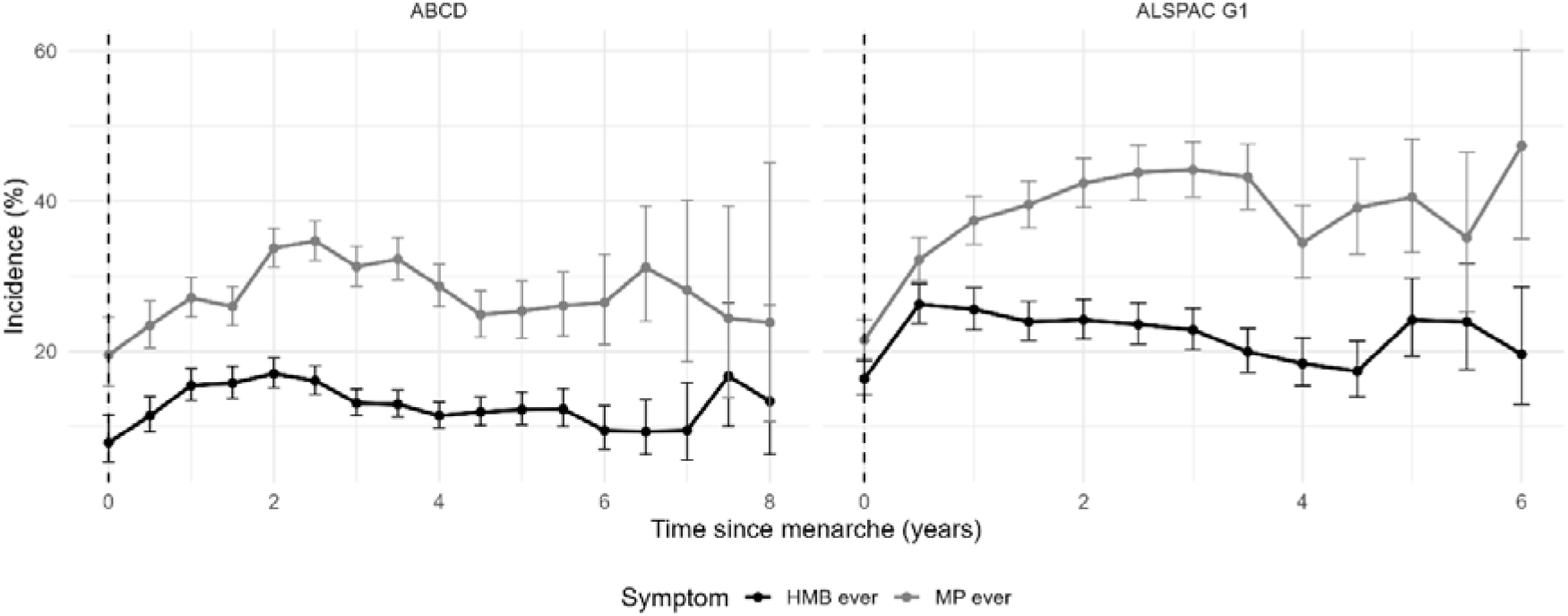
Age-specific incidence of heavy menstrual bleeding (HMB) and menstrual pain (MP) by time since menarche across adolescent and early adulthood cohorts.

## Discussion

In this multi-cohort study collectively spanning adolescence through to late reproductive life, we provide a harmonised life-course description of the prevalence and incidence of HMB and MP. We show that these symptoms are highly prevalent but exhibit distinct age- and sociodemographic-related patterns. MP predominates in adolescence, whereas HMB becomes increasingly prominent in mid (20-44 years) reproductive life and peaks in later (≥45 years) reproductive life.

Reports using other adolescent cohorts have also found a high prevalence of MP. In the Longitudinal Study of Australian Children, 35-50% of girls aged 14-18 reported MP ^22^. A systematic review and meta-analysis of 38 studies involving over 21,000 adolescents and young women estimated the pooled prevalence of dysmenorrhoea to be 71%, highlighting that menstrual pain is highly prevalent across diverse settings ^23^.

Our results are also consistent with a study of 14,247 participants from the Australian Longitudinal Study on Women’s Health, where the prevalence of HMB rose markedly from early adulthood (18% at age 22) to middle age (32% at age 48), whereas MP remains prevalent throughout but is especially dominant in younger participants ^2^. The higher prevalence of HMB with increasing age is supported by a Study of Women’s Health Across the Nation (SWAN) cohort study, which found that 35% of midlife women (aged 42-52) experienced three or more days of heavy bleeding ^24^.

Although we did not explore why these age-related changes in HMB and MP prevalence have been observed, they may be explained by parity as MP appears to decrease, whereas HMB may increase, with more child births ^27^. Also, HMB is often the result of underlying structural causes of abnormal uterine bleeding (which includes HMB), such as uterine fibroids, which become more common with age ^27^.

When examining the onset of HMB and MP during adolescence, we found that symptoms most commonly emerged within 1–3 years of menarche. Others have reported similar findings ^28^. This suggests that the first few years following menarche are an important window where the majority of menstrual symptoms begin, highlighting the need for education and support during this time to help manage such symptoms, particularly given the possible impacts of MP and HMB on attendance and attainment ^3^.

We also observed that individuals from lower socioeconomic backgrounds tended to have a higher prevalence of reporting either HMP or MP than those from more advantaged backgrounds across most of the cohorts. Generally, interpretation of ethnic differences was limited by low numbers of participants from minority ethnic groups, with prevalence estimates generally similar between groups. Although the strength of associations varied between cohorts, the overall pattern suggests that menstrual health inequalities may exist, consistent with previous studies. For example, financial difficulties and lower education have previously been associated with higher odds of painful and heavy periods in the two generations of ALSPAC ^29^. In the Reproductive Health Survey for England in 2023 (an online cross-sectional survey of over 59,000 women between 16 and 55 years old), financial insecurity was associated with higher odds of reporting painful and/or heavy periods ^30^. Inequalities likely exist due to broader structural determinants of health, such as access to nutritious food and safe spaces for physical activity, and dysregulation of hormonal and local endometrial inflammatory systems as a consequence of stress ^29^.

Key strengths of our study include the use of data from multiple large, population-based cohorts spanning adolescence through mid and late reproductive life enabling a unique life course perspective on menstrual symptoms. In addition, the longitudinal nature of the data allows for the examination of cumulative prevalence, point prevalence, and adolescent incidence, providing insights into symptom onset and progression.

Study limitations include reliance on self-report assessment of menstrual symptoms, which, while used clinically, may be subject to recall bias and differences in interpretation between participants and across cohorts. Although we attempted to harmonise definitions, variations in questionnaire items limits comparability and highlights the need for standardised menstrual symptom measures in cohort studies. Questionnaire measures also varied in reporting range, with some cohorts asking about most recent periods (e.g., ABCD and ALSPAC G0) and others asking about experiences *‘ever’* (e.g., ALSPAC G1). Longer reporting ranges may result in higher prevalence estimates and reporting error. No individual cohort covered the full reproductive life course, requiring life course patterns to be inferred by synthesising evidence across multiple cohorts with different age ranges, recruitment periods, and follow up durations. Additionally, all included cohorts were from HICs and predominantly from

White ethnic backgrounds, limiting the generalisability of our findings to LMICs, where menstrual experiences, access to healthcare and sociocultural contexts may differ. Finally, some sociodemographic indicators were not available across all cohorts, limiting the comparability of stratified analyses. To maximise comparability, we focused on childhood SEP; however, this may not fully reflect later socioeconomic circumstances at the time menstrual symptoms were reported in adult cohorts.

These findings have important implications for public health and clinical practice. The high prevalence of menstrual symptoms across the reproductive life course highlights the need for improved recognition and management at different life stages. While MP commonly emerges during adolescence and the early years following menarche, HMB is highly prevalent from mid to late reproductive life, highlighting the need for continued awareness and appropriate clinical assessment beyond adolescence. Schools and primary care settings may provide key opportunities for education, screening, and support during adolescence, while routine primary and gynaecological care should continue to recognise HMB throughout adulthood. Public health strategies to improve menstrual health should also address broader social and economic determinants that may contribute to menstrual health inequalities. Future research should focus on identifying risk factors for early onset and persistence of menstrual symptoms, as well as evaluating interventions to reduce the burden of menstrual symptoms across the life course. Longitudinal cohort studies with more detailed, standardised measures of menstrual symptoms and more ethnic diversity, will improve comparability across studies, enable more accurate estimates of symptom trajectories and risk factors, and strengthen the evidence base for advancing this field.

## Conclusion

HMB and MP are prevalent across the reproductive life course, with onset occurring predominantly in the early years following onset of menarche. MP is more prominent in early life, and HMB prevalence increases in later reproductive years. Sociodemographic differences suggest that menstrual health inequalities persist across populations, demonstrating that menstrual symptoms represent a substantial public health burden, yet they are often normalised and dismissed. Improved recognition and earlier intervention could be valuable in improving the management and treatment of HMB and MP across the life course. Ongoing and future cohort studies should endeavour to collect detailed, standardised measures of menstrual symptoms to enable comparison and better understanding of menstrual health across the life course, while including more diverse populations.

## Data Availability

The datasets analysed during the current study are available from the respective cohort studies subject to their data access policies and approval procedures. Data from the Adolescent Brain Cognitive Development (ABCD) Study are available through the National Institute of Mental Health Data Archive following an approved Data Access Request. ALSPAC data are available upon application to the ALSPAC Executive. Raine Study data are available upon application and approval by the Raine Study Executive Committee. MoBa data are available through the Norwegian Institute of Public Health subject to ethical and data access approvals. BCS70 data are available through the UK Data Service subject to registration and the relevant data access agreements.

https://abcdstudy.org/

http://www.bristol.ac.uk/alspac/researchers/our-data/

https://rainestudy.org.au/home-beta-2/information-for-researchers/project-application-process-sm/

https://www.helsedata.no

https://ukdataservice.ac.uk/

## Code availability

Statistical analysis and harmonisation code are publicly available on GitHub repository (https://github.com/sampurnakundu/MenstruLife.git). Harmonisation workflows and analytic scripts were independently reviewed by multiple authors to ensure reproducibility and consistency across cohorts.

## Contributions

SK, GS: conceptualisation, data curation, formal analysis, methodology, visualisation, writing – original draft, and writing – review & editing. GCS: conceptualisation, data curation, formal analysis, funding acquisition, investigation, methodology, project administration, resources, supervision, validation, visualisation, writing – review & editing. MCM: conceptualisation, data curation, formal analysis, supervision, validation, writing – review & editing. LDH, AH, MH, HOM, JT, JAM, HC, DJB: conceptualisation, supervision, writing – review & editing.

