## Supplementary information and figures for "Life-course patterns of heavy menstrual bleeding and menstrual pain across multiple population-based cohorts"

**Supplementary Material**

**STROBE Statement -** checklist of items that should be included in reports of observational studies

|  | Item No | Recommendation | Page |
| --- | --- | --- | --- |
| **Title and abstract** | 1 | (*a*) Indicate the study’s design with a commonly used term in the title or the abstract | 1 |
|  |  | (*b*) Provide in the abstract an informative and balanced summary of what was done and what was found | 2 |
| Introduction | | |  |
| Background/rationale | 2 | Explain the scientific background and rationale for the investigation being reported | 3-4 |
| Objectives | 3 | State specific objectives, including any prespecified hypotheses | 4 |
| Methods | | |  |
| Study design | 4 | Present key elements of study design early in the paper | 4 |
| Setting | 5 | Describe the setting, locations, and relevant dates, including periods of recruitment, exposure, follow-up, and data collection | 4 |
| Participants | 6 | (*a*) *Cohort study*—Give the eligibility criteria, and the sources and methods of selection of participants. Describe methods of follow-up  *Case-control study*—Give the eligibility criteria, and the sources and methods of case ascertainment and control selection. Give the rationale for the choice of cases and controls  *Cross-sectional study*—Give the eligibility criteria, and the sources and methods of selection of participants | 4, Fig S1 |
|  |  | (*b*) *Cohort study*—For matched studies, give matching criteria and number of exposed and unexposed  *Case-control study*—For matched studies, give matching criteria and the number of controls per case | - |
| Variables | 7 | Clearly define all outcomes, exposures, predictors, potential confounders, and effect modifiers. Give diagnostic criteria, if applicable | 5-6 |
| Data sources/ measurement | 8* | For each variable of interest, give sources of data and details of methods of assessment (measurement). Describe comparability of assessment methods if there is more than one group | 5-6, Table S1a-f, Table S2, Supplement File 1 |
| Bias | 9 | Describe any efforts to address potential sources of bias | 5-6 |
| Study size | 10 | Explain how the study size was arrived at | 4, Fig S1 |
| Quantitative variables | 11 | Explain how quantitative variables were handled in the analyses. If applicable, describe which groupings were chosen and why | 5-6 |
| Statistical methods | 12 | (*a*) Describe all statistical methods, including those used to control for confounding | 6 |
|  |  | (*b*) Describe any methods used to examine subgroups and interactions | 6 |
|  |  | (*c*) Explain how missing data were addressed | - |
|  |  | (*d*) *Cohort study*—If applicable, explain how loss to follow-up was addressed  *Case-control study*—If applicable, explain how matching of cases and controls was addressed  *Cross-sectional study*—If applicable, describe analytical methods taking account of sampling strategy | - |
|  |  | (*e*) Describe any sensitivity analyses | - |
| **Results** |  |  |  |
| Participants | 13* | (a) Report numbers of individuals at each stage of study—eg numbers potentially eligible, examined for eligibility, confirmed eligible, included in the study, completing follow-up, and analysed | 4, Fig S1 |
|  |  | (b) Give reasons for non-participation at each stage | Fig S1 |
|  |  | (c) Consider use of a flow diagram | Fig S1 |
| Descriptive data | 14* | (a) Give characteristics of study participants (eg demographic, clinical, social) and information on exposures and potential confounders | 7, Table 2 |
|  |  | (b) Indicate number of participants with missing data for each variable of interest | Fig S1, Table 2 |
|  |  | (c) *Cohort study*—Summarise follow-up time (eg, average and total amount) | 4, Supplement File 1 |
| Outcome data | 15* | *Cohort study*—Report numbers of outcome events or summary measures over time | 7-8 |
|  |  | *Case-control study—*Report numbers in each exposure category, or summary measures of exposure | - |
|  |  | *Cross-sectional study—*Report numbers of outcome events or summary measures | - |
| Main results | 16 | (*a*) Give unadjusted estimates and, if applicable, confounder-adjusted estimates and their precision (eg, 95% confidence interval). Make clear which confounders were adjusted for and why they were included | 7-9, Tables S3, S4, S5, S6 |
|  |  | (*b*) Report category boundaries when continuous variables were categorized | - |
|  |  | (*c*) If relevant, consider translating estimates of relative risk into absolute risk for a meaningful time period | - |
| Other analyses | 17 | Report other analyses done—eg analyses of subgroups and interactions, and sensitivity analyses | - |
| **Discussion** |  |  |  |
| Key results | 18 | Summarise key results with reference to study objectives | 9 |
| Limitations | 19 | Discuss limitations of the study, taking into account sources of potential bias or imprecision. Discuss both direction and magnitude of any potential bias | 10 |
| Interpretation | 20 | Give a cautious overall interpretation of results considering objectives, limitations, multiplicity of analyses, results from similar studies, and other relevant evidence | 9-11 |
| Generalisability | 21 | Discuss the generalisability (external validity) of the study results | 10 |
| **Other information** |  |  |  |
| Funding | 22 | Give the source of funding and the role of the funders for the present study and, if applicable, for the original study on which the present article is based | 11 |

**Note:** An Explanation and Elaboration article discusses each checklist item and gives methodological background and published examples of transparent reporting. The STROBE checklist is best used in conjunction with this article (freely available on the Web sites of PLoS Medicine at http://www.plosmedicine.org/, Annals of Internal Medicine at http://www.annals.org/, and Epidemiology at http://www.epidem.com/). Information on the STROBE Initiative is available at www.strobe-statement.org.

**Cohorts**

**Adolescent Brain Cognitive Development (ABCD)**

*Description:* The ABCD Study is an ongoing prospective longitudinal study starting at the ages of 9-10 and following participants for 10 years. The study includes a diverse sample of 11,878 children between 2016 and 2018 youth enrolled at 21 research sites across the United States. It measures brain development (via structural, task functional, and resting state functional imaging), social, emotional, and cognitive development, mental health, substance use and attitudes, gender identity and sexual health, biospecimens, as well as a variety of physical health, and environmental factors. In addition, various external databases have been linked with ABCD Study data providing information about local conditions for environment, poverty, pollution, school, and policy, as examples. These can be used for providing context when evaluating behavioral and brain development ^1,2^. For more information about the ABCD Study, please visit the ABCD Study website: <https://abcdstudy.org/>.

*Acknowledgments and funding:* We thank the ABCD Study participants and families, as well as the ABCD research staff and consortium investigators, for making these data available. The ABCD Study is supported by NIH partners (the National Institute on Drug Abuse, the National Institute on Alcohol Abuse and Alcoholism, the National Cancer Institute, the Eunice Kennedy Shriver National Institute of Child Health and Human Development, the National Institute of Mental Health, the National Institute on Minority Health and Health Disparities, the National Institute of Neurological Disorders and Stroke, the National Heart, Lung, and Blood Institute, the National Institute of Justice, the NIH Office of Behavioral and Social Sciences Research, and the NIH Office of Research on Women’s Health), as well as the Centers for Disease Control and Prevention – Division of Adolescent and School Health.

*Data availability:* Data from the Adolescent Brain Cognitive Development (ABCD) Study are available through the National Institute of Mental Health (NIMH) Data Archive (https://nda.nih.gov/abcd), subject to an approved Data Access Request (DAR).

*Ethics:* Most ABCD research sites rely on a central Institutional Review Board (cIRB) at the University of California, San Diego for the ethical review and approval of the research protocol, with a few sites obtaining local IRB approval ^3^.

**Avon Longitudinal Study of Parents and Children (ALSPAC)**

*Description:* Pregnant women resident in Avon, UK with expected dates of delivery between 1st April 1991 and 31st December 1992 were invited to take part in the study ^4,5^. The initial number of pregnancies enrolled was 14,541, resulting in 13,988 children who were alive at 1 year of age. Of the original 14,541 initial pregnancies, 338 were from a woman who had already enrolled with a previous pregnancy, meaning 14,203 unique mothers were initially enrolled in the study. When the oldest children were approximately 7 years of age, an attempt was made to bolster the initial sample with eligible cases who had failed to join the study originally. As a result, when considering variables collected from the age of seven onwards, there are data available for 15,447 pregnancies, resulting in 14,901 children were alive at 1 year of age (Generation 1; G1 offspring) and 14,833 unique women (Generation 0; G0 mothers). G0 mothers and G1 offspring have been regularly followed up through questionnaires and clinic visits. Please note that the study website contains details of all the data that is available through a fully searchable data dictionary and variable search tool and reference the following webpage: <http://www.bristol.ac.uk/alspac/researchers/our-data/>.

*Acknowledgements:* We are extremely grateful to all the families who took part in this study, the midwives for their help in recruiting them, and the whole ALSPAC team, which includes data collection staff, data and administrations staff, technical managers and the technical staff with the Bristol Bioresource Laboratory, based within the University of Bristol.

*Funding:* The UK Medical Research Council and Wellcome (Grant ref: MR/Z505924/1) and the University of Bristol provide core support for ALSPAC. This publication is the work of the authors and SK, GS, and GCS will serve as guarantors for the contents of this paper. A comprehensive list of grants funding is available on the ALSPAC website (<http://www.bristol.ac.uk/alspac/external/documents/grant-acknowledgements.pdf>).

*Data availability:* The informed consent obtained from ALSPAC (Avon Longitudinal Study of Parents and Children) participants does not allow the data to be made available through any third party maintained public repository. Supporting data are available from ALSPAC on request under the approved proposal number, B4396. Full instructions for applying for data access can be found here: http://www.bristol.ac.uk/alspac/researchers/access/. The ALSPAC study website contains details of all available data: <http://www.bristol.ac.uk/alspac/researchers/our-data/>.

*Ethics:* Ethical approval for the study was obtained from the ALSPAC Ethics and Law Committee and the Local Research Ethics Committees. Informed consent for the use of all data collected was obtained from participants following the recommendations of the ALSPAC Ethics and Law Committee at the time. Participants can contact the study team at any time to retrospectively withdraw consent for their data to be used. Study participation is voluntary and during all data collection sweeps, information was provided on the intended use of data. The completion of a questionnaire, either on paper or online, was considered to be written consent from participants to use their data for research purposes.

**The Raine Study**

*Description:* The Raine Study (previously referred to as the Western Australian Pregnancy Cohort) is an ongoing longitudinal, population-based, multigenerational birth cohort established in Perth, Western Australia. A total of 2,868 live births from 2,900 pregnancies were recruited between May 1989 and November 1991 from King Edward Memorial Hospital and surrounding clinics, with participants enrolled during pregnancy at approximately 18 weeks’ gestation. The children (Gen2) have been followed prospectively from pregnancy through childhood, adolescence, and adulthood with repeated assessments of health and development ^6–8^.

*Acknowledgements and Funding:* We gratefully acknowledge all the Raine Study participants and their families for their continued participation in the study, as well as the Raine Study team for study co-ordination and data collection. We also thank the NHMRC and the Raine Medical Research Foundation for their support. The core management of the Raine Study is funded by The University of Western Australia, Curtin University, The Kids Research Institute Australia, Women and Infants Research Foundation, Edith Cowan University, Murdoch University, The University of Notre Dame Australia and the Western Australian Future Health Research and Innovation Fund (Grant ID WACSOSP2023-2024, WACSOSP2025/7). Then Gen2-14 year follow-up was funded by NHMRC grants (Sly et al., 211912 and Stanley et al., 003209) and The Raine Medical Research Foundation. The Gen2-27 year follow up was funded by NHMRC grants (Sly et al., 211912 and Eastwood et al., 1109057), the National Heart Foundation (Mori et al., 102301), Cancer Australia (Stone et al., 1147677), Royal Perth Hospital Medical Research Foundation, The Heart Foundation (WA Branch), Lions Eye Institute, Curtin University, School of Population and Global Health (The University of Western Australia), Division of Obstetrics and Gynaecology (The University of Western Australia), and John Olynyk.

*Data availability:* The Raine Study data is available to researchers through project application and approval by the Raine Study Executive Committee: <https://rainestudy.org.au/home-beta-2/information-for-researchers/project-application-process-sm/>.

*Ethics:* Ethics approval for the Raine Study was obtained from the relevant Human Research Ethics Committees (2025/ET000219), and all participants provided written informed consent.

**Norwegian Mother, Father, and Child Cohort Study (MoBa)**

*Description:* The Norwegian Mother, Father and Child Cohort Study (MoBa) is a population-based pregnancy cohort study conducted by the Norwegian Institute of Public Health. Participants were recruited from all over Norway from 1999-2008. The women consented to participation in 41% of the pregnancies. The cohort includes approximately 114,500 children, 95,200 mothers and 75,200 fathers. MoBa is regulated by the Norwegian Health Registry Act. Data collection includes questionnaire-based information collected during pregnancy and at multiple follow-up timepoints, alongside linkage to national health registries. The cohort provides extensive data on maternal and offspring health, lifestyle, and socio-demographic factors ^9,10^. The current study was approved by The Regional Committees for Medical and Health Research Ethics of South/East Norway (reference number: 277291).

*Acknowledgements and funding:* We are grateful to all the participating families in Norway who take part in this on-going cohort study. We thank the Norwegian Institute of Public Health for generating high-quality data and all researchers and data scientists who contribute to enriching the data. The Norwegian Mother, Father and Child Cohort Study is supported by the Norwegian Ministry of Health and Care Services and the Ministry of Education and Research.

*Data availability:* Data from the Norwegian Mother, Father and Child Cohort Study is managed by the Norwegian Institute of Public Health. Access requires approval from the Regional Committees for Medical and Health Research Ethics (REC), compliance with GDPR, and data owner approval. Participant consent does not allow individual-level data storage in repositories or journals. Researchers seeking access for replication must apply via [www.helsedata.no](http://www.helsedata.no).

*Ethics:* The establishment of MoBa and initial data collection were based on a license from the Norwegian Data Protection Agency and approval from the Regional Committees for Medical and Health Research Ethics. The MoBa cohort is currently regulated by the Norwegian Health Registry Act. Each new research project needs to obtain ethical approval for using the data collected in MoBa for the specific research questions of interest.

**1970 British Cohort Study (BCS70)**

*Description:* The 1970 British Cohort Study (BCS70) is a nationally representative longitudinal birth cohort study in the United Kingdom that follows individuals born in a single week of April 1970. The original cohort comprised 17,198 births in England, Scotland, and Wales, with subsequent follow-up sweeps conducted across childhood and adulthood, collecting information on health, social, and economic outcomes ^11^.

*Acknowledgements and funding:* BCS70 is funded by the Economic and Social Research Council and managed by the UCL Centre for Longitudinal Studies.

*Data availability:* Data from BCS70 are held and distributed by the UK Data Archive, a service provider of the Economic and Social Data Service. Non-commercial users can download data free of charge, subject to registration and data access agreements (<https://ukdataservice.ac.uk/>). Further information about the cohort and other longitudinal studies can be obtained from ESDS longitudinal at <http://www.esds.ac.uk/longitudinal/>.

*Ethics:* Multicentre Research Ethics Committee (MREC) ethical approval has been sought for BCS70 follow-ups from 2000 onwards. The 1970, 1975, 1980, 1986 and 1996 follow-ups pre-dated the establishment of the MREC system. Available records suggest that there was only internal ethical review for these surveys

**Age**

In ALSPAC G0, individual participant age at assessment was used to define timepoint age for calculating age-specific point prevalence. In ABCD and ALSPAC G1, individual participant age at assessment was used to examine symptom onset (incidence) with chronological age.

Age was rounded down to the nearest year in all cohorts. In instances where less than 40 participants had provided data at a certain age, age years were grouped (i.e., ≥56 years in ALSPAC G0 and 9/10 and 11/12 in ALSPAC G1). We also excluded responses at age 18 and 19 in ALSPAC G0 due to very small observation numbers and to avoid misrepresenting ALSPAC G0 as an adolescent cohort. Also, where individual participant age at assessment was utilised, there were a small number of instances (<3% of all observations) where participants had completed subsequent questionnaires within less than 12 months of each other, resulting in two responses at the same age. To manage these duplicates, we defined a participant as experiencing HMB and/or MP if they reported this symptom at either of the responses at the same age.

**Gynaecological age**

In ABCD and ALSPAC G1, we examined incidence with gynaecological age (or time since menarche), which was calculated by subtracting age at menarche from age at assessment (as above; prior to being rounded down) ^12,13^. Time since menarche was rounded down to the nearest six months. As with age, gynaecological age groups were combined where there were less than 40 observations (≥8 years in ABCD) and any duplicates were handled by defining participant as experiencing HMB and/or MP if they reported this symptom at either of the responses at the same 6-month gynaecological age band.

**Supplementary Figures**

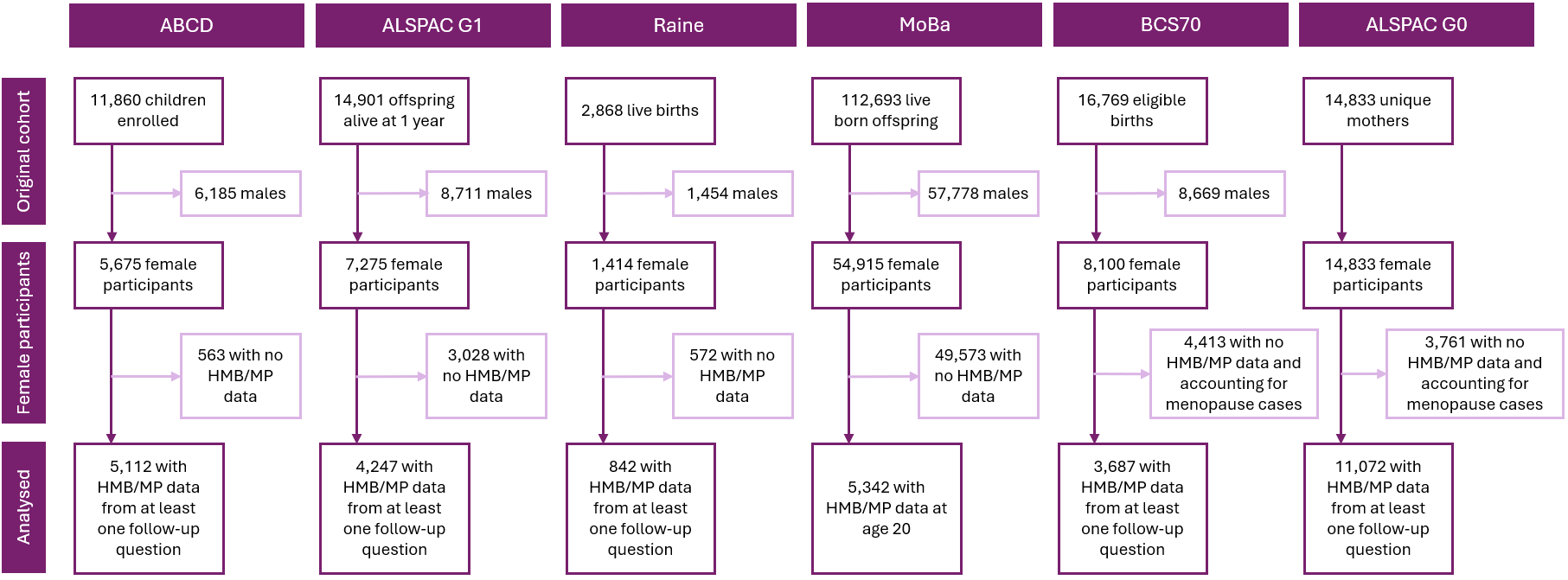

**Figure S1:** Cohort-wise sample breakdown flowchart

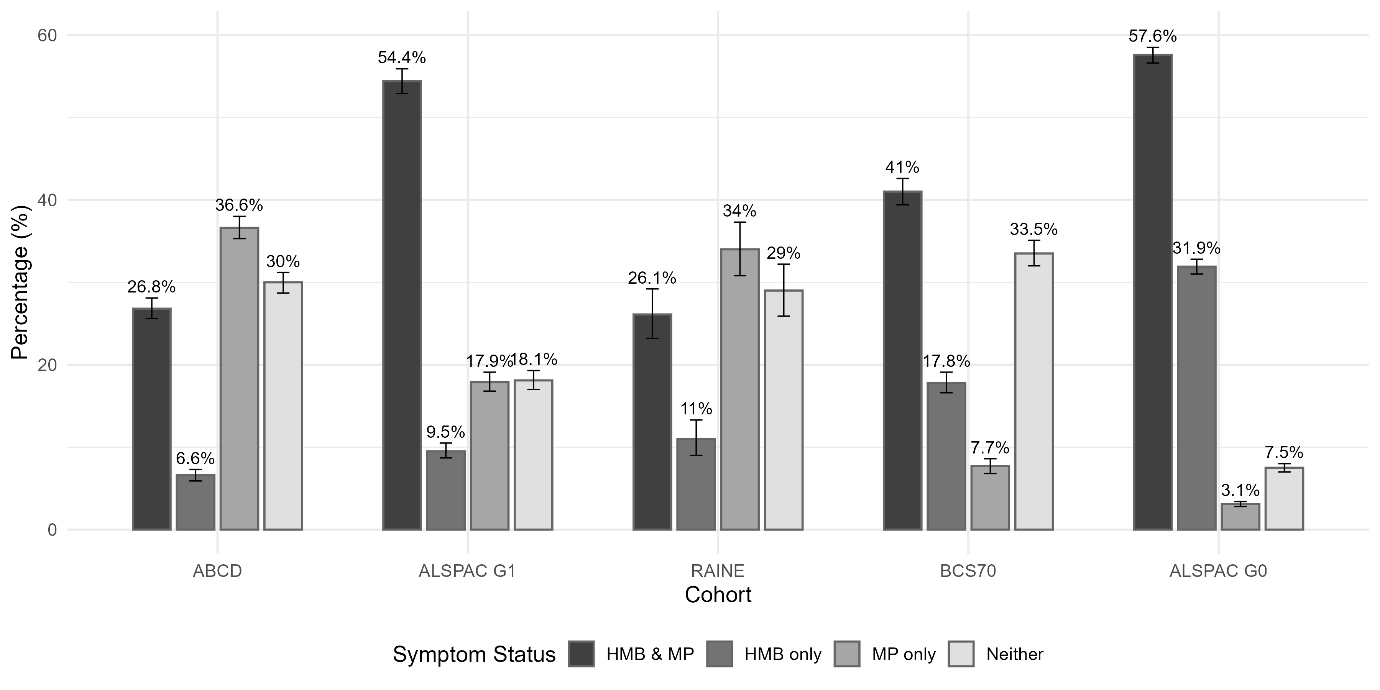

**Figure S2:** Cumulative prevalence of HMB only, MP only, Co-occurrence and Neither symptoms

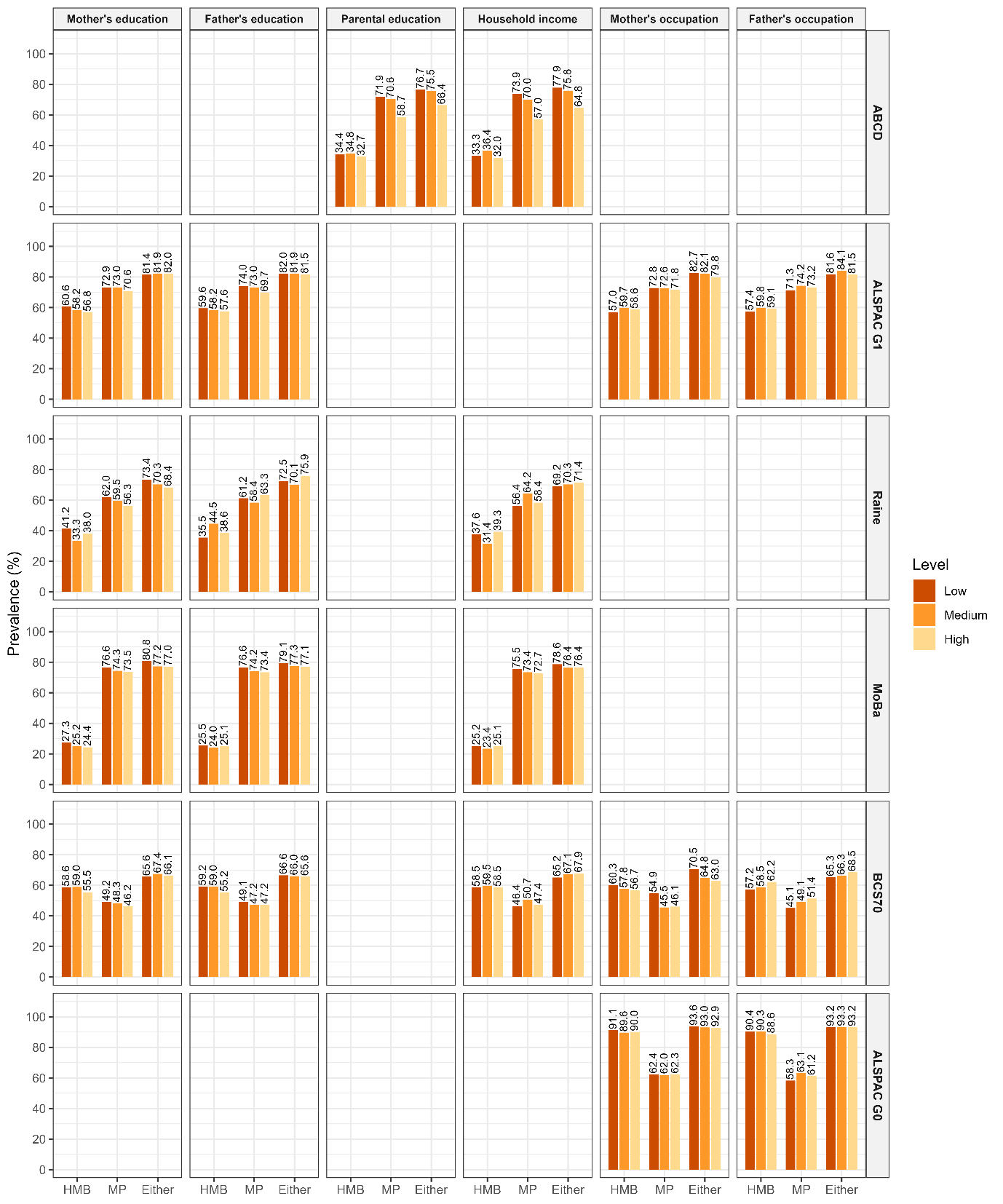

**Figure S3**: Prevalence of HMB, MP, and either symptom by socioeconomic position across cohorts.

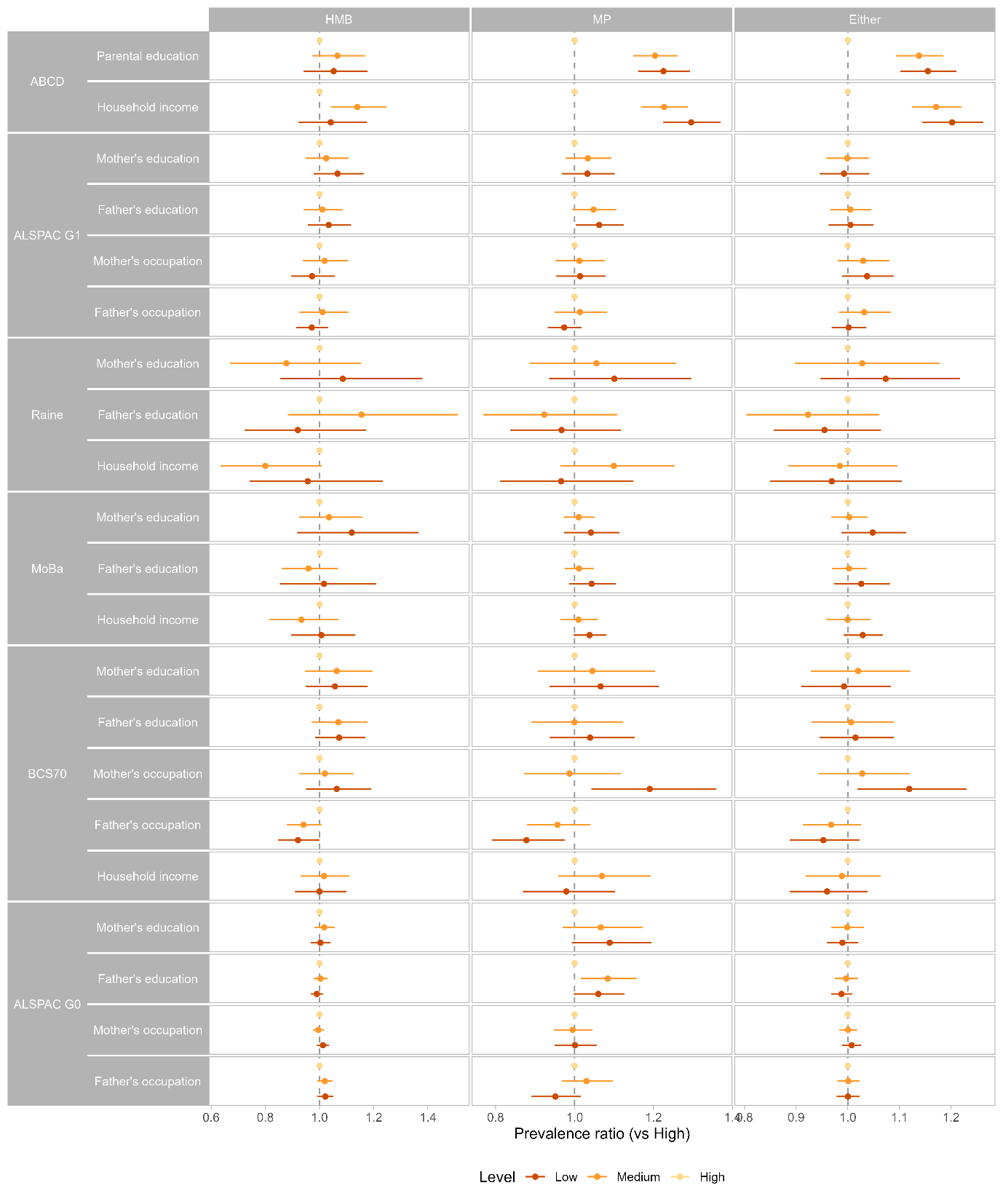

**Figure S4:** Prevalence ratios for HMB, MP, and either symptom according to socioeconomic position across cohorts.

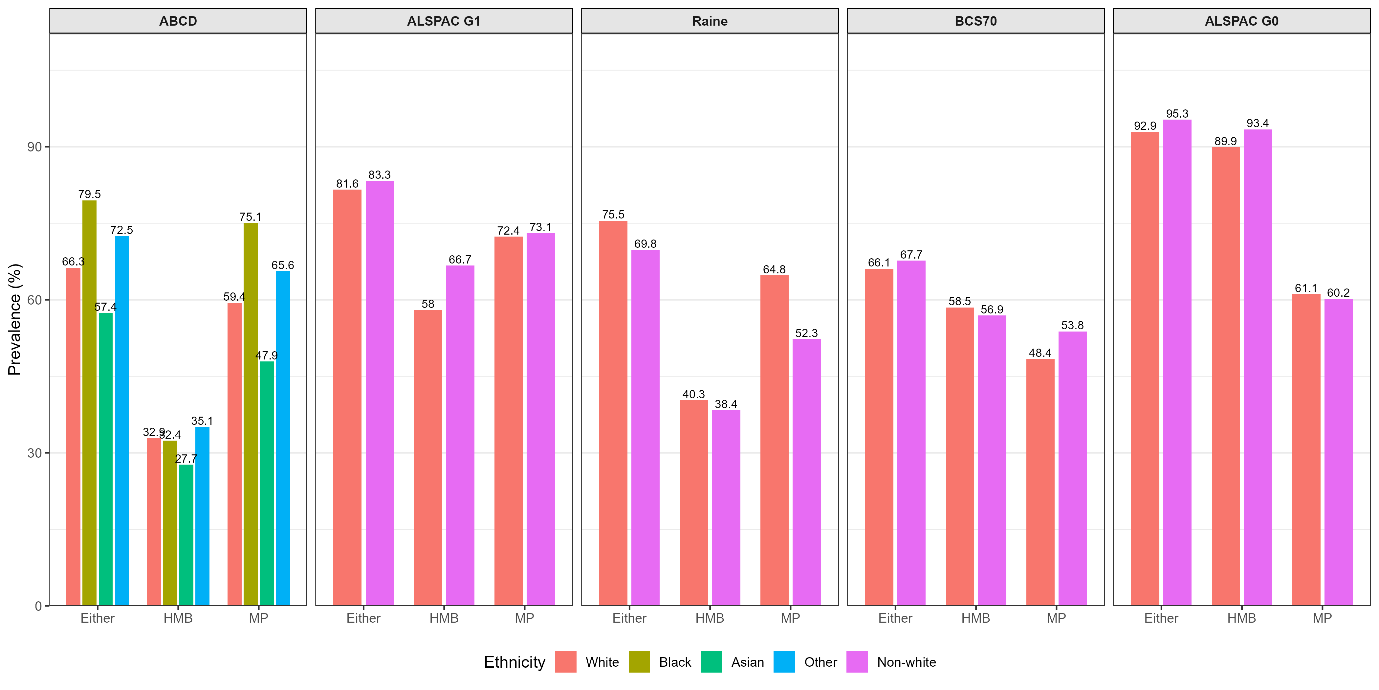

**Figure S5**: Prevalence of HMB, MP and either symptom by ethnicity across cohorts
